# The Measuring Advanced Practitioner Presence, Patterns, and Evolving Distribution (MAPPED) study: Protocol for secondary data analysis

**DOI:** 10.64898/2026.06.27.26356732

**Authors:** Sadie Diamond-Fox, Barry Hill

**Affiliations:** School of Healthcare and Nursing Sciences, Faculty of Health and Wellbeing, Northumbria University, Coach Lane Campus (East), Newcastle upon Tyne, United Kingdom

## Abstract

**Background:** Advanced Practitioners (APs) are a growing multi-professional workforce within the National Health Service (NHS) in England, spanning nursing, pharmacy, allied health, and healthcare science disciplines. Despite their policy prominence, no national quantification of this workforce using routinely collected administrative data has been published. Existing intelligence relies on regional surveys, employer self-reports, and bespoke data requests, each limited in coverage and comparability. This protocol describes a secondary analysis of NHS Workforce Statistics to address this gap, to support national workforce planning and service redesign.

**Methods:** The Measuring Advanced Practitioner Presence, Patterns, and Evolving Distribution (MAPPED) study is a secondary analysis of NHS Workforce Statistics derived from the Electronic Staff Record (ESR), examining the AP workforce in Hospital and Community Health Services (HCHS) in England. The study combines longitudinal analysis of national trends from September 2014 to April 2026 (13 time points) with cross-sectional geographic analysis at NHS England region and Integrated Care System (ICS) levels in April 2026. APs are identified using National Workforce Dataset Job Role codes. The analytical framework comprises descriptive statistics, Mann-Kendall trend tests, Kruskal-Wallis tests, Gini coefficients, and Wilson score confidence intervals; trainee Advanced Practitioners are analysed separately. Reporting follows the REporting of studies Conducted using Observational Routinely-collected health Data (RECORD) statement and the STandardisierte BerichtsROutine für SekundärdatenAnalysen 2 (STROSA-2) checklist for secondary data analyses.

**Results and analysis:** This paper presents the study protocol; no results are reported. Planned outputs include national trend figures, distribution tables by staff group and Area of Work, geographic inequality measures using Lorenz curves, and sensitivity analyses addressing classification-version effects and generic code persistence.

**Discussion:** This study will provide the first national, longitudinal quantification of the AP workforce from routinely collected administrative data. The principal limitation is that Job Role coding accuracy varies across trusts, and the transition to profession-specific codes from the year 2022 creates a measurement discontinuity that sensitivity analyses address but cannot fully eliminate.

## Introduction

Advanced Practitioner (AP) roles have become a prominent feature of workforce redesign within the National Health Service (NHS) Hospital and Community Health Services (HCHS) in England. These roles span multiple registered professions, including nursing, pharmacy, paramedicine, physiotherapy, and other allied health and healthcare science disciplines. The Multi-Professional Framework for Advanced Practice in England (MPF), first published in 2017 [1] and updated in 2025 [2], defines APs as clinicians educated to Master’s level or equivalent who demonstrate autonomous, complex decision-making across four pillars of practice: clinical practice, leadership and management, education, and research. APs are deployed across a broad range of clinical settings, from emergency departments and acute medicine to community and mental health services [2,3]. The MPF provides the national definition of advanced clinical practice against which the National Workforce Data Set (NWD) Job Role codes used in this study were developed; the profession-specific AP codes introduced from NWD v3.3 onwards were designed to enable identification of staff working at the level described in the MPF within the NHS Electronic Staff Record (ESR).

The NHS Long Term Workforce Plan identifies advanced practice as a mechanism for increasing capacity in specialist services and supporting career development pathways [3]. Despite this policy commitment, the evidence base for understanding the AP workforce at national level is fragmented. To date, no national longitudinal analysis of the AP workforce using routinely collected NHS administrative data has been published. The routine workforce statistics published by NHS England do not contain a discrete “Advanced Practitioner” reporting category in their standard outputs. Identifying APs requires cross-referencing NWD Job Role codes, which were only introduced in profession-specific form from February 2022, with NWD Staff Group classifications [4]. Large national surveys have documented considerable variation in role titles, scope of practice, job descriptions, and educational backgrounds [5,6]. Qualitative and mixed-methods studies have identified high levels of ambiguity in how the role is understood and deployed, with persistent challenges around governance, supervision, and professional identity [7–9]. A UK-wide scoping review found that the vast majority of evidence focused on nursing, pharmacy, physiotherapy, and radiography, with minimal coverage of other professions and settings [10].

Workforce planning, service redesign, and the equitable deployment of advanced practice capacity all depend on reliable national intelligence: where Advanced Practitioners work, which professions they come from, how coding systems represent them, and where future supply April emerge. Without it, these decisions rest on partial and self-reported data. The global health workforce crisis has intensified pressure on high-income health systems to reconfigure professional roles and skill mix [11,12]. The World Health Organization’s Global Strategy on Human Resources for Health: Workforce 2030 called for optimising the existing workforce through task sharing and expanded scopes of practice [13]. A five-year review of the Strategy’s implementation found that while progress had been made in data collection and information exchange, many countries remained underprepared for the workforce demands exposed by the COVID-19 pandemic, and that reliable national workforce intelligence remained a prerequisite for effective policy responses [14]. These findings reinforce the case for granular, routinely derived workforce data of the kind this study seeks to produce.

International evidence demonstrates that advanced-practice workforce growth can be rapid and geographically uneven, with supply varying substantially by region and regulatory context [15,16]. The United States offers one well-documented example: the share of Medicare evaluation and management visits delivered by nurse practitioners and physician assistants increased from 14.0% to 25.6% between 2013 and 2019 [17]. Similar variation in advanced-practice supply and role development appears across other high-income systems [18,19]. Most of this evidence sits outside England; whether the same patterns hold for the NHS advanced-practice workforce is not yet established. Geographic workforce intelligence bears directly on access and equity.

Quantifying the AP workforce from administrative data presents specific methodological challenges. The Electronic Staff Record (ESR), the integrated human resources and payroll system used by NHS organisations in England, is the primary source of national workforce statistics. ESR contains NWD Job Role codes that can identify Advanced Practitioners, but the accuracy of these codes depends on employer coding practice, which varies across organisations [4,20]. Before February 2022, only generic “Advanced Practitioner” codes existed within the NWD, without profession-specific disaggregation. The introduction of profession-specific codes from NWD version 3.3 onwards represented a substantial improvement in classification granularity, but created challenges for interpreting longitudinal trends: apparent changes in professional composition April reflect coding reclassification rather than genuine workforce shifts. Regional workforce reports have noted that coding adoption is incomplete, with some trusts retaining staff on the older generic codes [21]. These data quality considerations mean that any national analysis must be transparent about the identification strategy, acknowledge coding-transition effects, and include sensitivity analyses to test the influence of classification changes on the findings.

Internationally, the difficulties of AP workforce enumeration are well documented. A global survey of 325 respondents across 26 countries reported that understanding the number, distribution, and types of advanced practice providers is “extremely complicated” because of categorisation issues specific to nursing and advanced practice [22]. A six-country analysis of nurse practitioner workforce size excluded UK nations because no suitable national registry or data source was identified [18]. An Organisation for Economic Co-operation and Development (OECD) review of advanced practice nursing in 12 developed countries found wide variation in role definitions, educational requirements, and regulatory frameworks, and recommended that countries invest in standardised data collection [19]. In Australia, a national nursing workforce survey of 5,662 respondents found that position titles alone were insufficient for classification and developed a role-delineation tool to identify practitioners working at an advanced level [23]. In the United States, a validation study of the National Provider Identifier file found that it undercounted advanced practice midwives because multiple taxonomy categories existed for the same role [24]. The data quality challenges facing this study are common to advanced practice workforce enumeration internationally.

A secondary analysis of routinely collected NHS Workforce Statistics is the appropriate study design for this research question. The NHS Workforce Statistics provide a near-complete census of HCHS staff at each publication time point, covering all NHS Trusts and core organisations in England. No primary data collection method, whether survey, audit, or registration-based, could achieve equivalent national coverage at comparable cost or with the longitudinal depth available in the published time series. Survey-based approaches, while valuable for capturing scope of practice and qualitative workforce characteristics, are subject to response bias and cannot produce the population-level denominators needed for workforce planning. The publicly available NHS Workforce Statistics, published under the Open Government Licence v3.0, provide a reproducible data source that other researchers can access and verify.

## Study aim and objectives

The aim of this study is to provide the first national quantification of the Advanced Practitioner workforce within NHS Hospital and Community Health Services in England, using routinely collected workforce data to examine workforce composition, growth, clinical deployment, and geographic distribution.

The study addresses five objectives:

1. To describe the size and growth trajectory of the qualified AP workforce (full-time equivalent (FTE)) at national level across the study period (September 2014 to April 2026), overall and by NWD Staff Group.
2. To examine the distribution of qualified APs across Areas of Work (clinical specialties) and how this distribution has changed over time.
3. To characterise the transition from generic to profession-specific Advanced Practitioner Job Role codes following the NWD v3.3 classification changes, including the persistence of generic coding and the progressive introduction of new profession-specific codes across NWD versions 3.3 to 3.6.
4. To examine the geographic distribution of qualified AP FTE across NHS England regions and Integrated Care Systems (ICS) at April 2026, including the degree of geographic inequality in workforce deployment.
5. To describe the size and distribution of the Trainee AP workforce as a supplementary analysis, providing an indication of the future AP workforce pipeline.

All objectives are descriptive and non-causal. The study characterises distributional properties and detects trends; it does not test hypotheses about determinants of AP workforce size or distribution. The methodological framing for this approach is set out in the Study design and reporting framework section below.

## Materials and methods

### Study design and reporting framework

The MAPPED study is a pre-specified, observational, descriptive secondary analysis of routinely collected NHS workforce administrative data. The analytical framework is descriptive and non-causal, conducted within the descriptive epidemiology framework articulated by Lesko et al. [25], which requires pre-specification of the target population, the outcome, the measure of occurrence, and the role of any auxiliary variables, and within the ’description’ task category defined by Hernán, Hsu and Healy [26], which distinguishes description from prediction and causal inference as a legitimate analytical goal in its own right. The approach is consistent with the conventions of major workforce reporting bodies, including the Organisation for Economic Co-operation and Development (OECD) [27], the World Health Organization (WHO) [13], the Health Foundation [28], and the Nuffield Trust [29,30]. No experimental manipulation, randomisation, or blinding is involved. The study does not test causal hypotheses, does not estimate causal effects, and does not employ regression models or adjusted analyses.

The study combines two complementary analytical components. The first is a longitudinal repeated cross-sectional analysis of national AP workforce trends using Core 7 of the NHS Workforce Statistics (September 2014 to April 2026, 13 time points). The second is a cross-sectional analysis of geographic distribution using Core 8 (April 2026 only). The design is repeated cross-sectional rather than panel: Core 7 reports aggregate national full-time equivalent (FTE) values at each snapshot, not individual-level trajectories tracked over time. This distinction matters because the study characterises workforce patterns at successive time points but cannot attribute changes to the movement of specific individuals between organisations or roles.

This study is reported in accordance with the REporting of studies Conducted using Observational Routinely-collected health Data (RECORD) statement, an extension of the Strengthening the Reporting of OBservational studies in Epidemiology (STROBE) guidelines for observational studies using routinely collected health data [31]. RECORD was selected as the primary reporting guideline because the study analyses administrative workforce data collected for operational purposes by NHS England, which falls within RECORD’s defined scope. RECORD does not, however, set out as explicit items a small number of reporting dimensions that are relevant to secondary analyses of administrative data. For these, supplementary items were drawn from the STandardisierte BerichtsROutine für SekundärdatenAnalysen, version 2 (STROSA-2) checklist [32]: legal basis for data use (item 7), data flow (item 9), unit of analysis (item 11), strengths and weaknesses of the secondary data approach (item 21), and role of the data owner (item 26). STROSA-2 is a national reporting standard developed for secondary data analyses in Germany; only those items applicable to this study and not already covered by RECORD were adopted, rather than the full 27-item checklist. The completed RECORD checklist (S1 Checklist) and a table mapping each supplementary STROSA-2 item to its location in the manuscript (S2 Table) are provided as supporting information.

### Data source and provenance

The primary data source is the NHS Workforce Statistics publication series, produced by NHS England (formerly NHS Digital). This series reports monthly numbers of Hospital and Community Health Services (HCHS) staff working in NHS Trusts and other core organisations in England, excluding primary care staff. Data are derived from validated extracts of the Electronic Staff Record (ESR), the integrated human resources and payroll system used by NHS organisations across England. NHS Workforce Statistics are classified as Official Statistics under the Statistics and Registration Service Act 2007 and are published under the Open Government Licence v3.0, which permits secondary analysis and publication of derived findings without requiring ethical approval or data access agreements [20]. The March 2026 NHS Workforce Statistics release was downloaded from the NHS England NHS Workforce Statistics publication page on 1 April 2026 for preliminary structural inspection. The analytical dataset, the April 2026 release, will be downloaded from the same source following its scheduled publication on 25 June 2026. The legal basis for data access and analysis is the Open Government Licence v3.0, under which NHS Workforce Statistics are published. No data access agreement, data sharing contract, or approval from the data owner (NHS England) was required, as the data are freely available for secondary analysis and publication of derived findings under the terms of the licence (STROSA-2 item 7). NHS England, as the owner of the source data, had no role in the design, conduct, analysis, or interpretation of this study, and the authors are solely responsible for the content of this manuscript and the decision to publish (STROSA-2 item 26).

Two data files form the primary analytical dataset. Both use the NWD classification system, which is the only classification in the published NHS Workforce Statistics that contains AP-specific Job Role codes. Table 1 summarises the structure and scope of each file.

**Table 1.** Primary analytical data files.

| Feature | Core 7 | Core 8 |
| --- | --- | --- |
| **Content** | FTE by NWD Staff Group, Job Role, and Area of Work at national level | FTE by NWD Staff Group, Job Role, and Area of Work at organisation level |
| **Analytical role** | Longitudinal analysis | Geographic distribution analysis |
| **Temporal coverage** | Annual September snapshots (September 2009 to September 2025) plus April 2026 endpoint | Single time point (April 2026) |
| **Column structure** | DATA_MONTH<br>DATA_LEVEL<br>NWD_STAFF_GROUP<br>NWD_JOB_ROLE<br>PRIMARY_AREA_OF_WORK<br>SECONDARY_AREA_OF_WORK<br>TERTIARY_AREA_OF_WORK<br>FTE | DATA_MONTH<br>DATA_LEVEL<br>NHSE_REGION_CODE<br>NHSE_REGION_NAME<br>ICS_CODE<br>ICS_NAME<br>ORG_CODE<br>ORG_NAME<br>NWD_STAFF_GROUP<br>NWD_JOB_ROLE<br>PRIMARY_AREA_OF_WORK<br>SECONDARY_AREA_OF_WORK<br>TERTIARY_AREA_OF_WORK<br>FTE |
| **Geographic identifiers** | None (national aggregate only) | NHS England region code and name, ICS code and name, organisation code and name |

Core 8 includes embedded region and ICS identifiers alongside organisation codes, which permits direct aggregation to region and ICS level without requiring an external reference table. Historical Core 8 files with the NWD classification are not published by NHS England for earlier time points, so the geographic analysis is limited to a single cross-sectional snapshot.

An important structural feature of the NHS Workforce Statistics is that the downloadable CSV files have changed format over time. The standard comma-separated values (CSV) package for earlier publications contains files structured around the main staff group classification derived from NHS Occupation Codes. These files do not contain NWD Staff Group or Job Role variables and cannot be used to identify Advanced Practitioners. The Core 7 and Core 8 files using the NWD classification are a separate data product. This data availability constraint is the reason the longitudinal analysis operates at national level only (using Core 7), while the geographic analysis is cross-sectional (using the April 2026 Core 8). Fig 1 illustrates the data flow from the ESR through the published data files to the analytical datasets used in this study, in accordance with STROSA-2 item 9.

**Fig 1. Data flow from the Electronic Staff Record to the analytical datasets.**

Supplementary data files from the standard CSV publication format April be used to provide contextual workforce denominators (total HCHS FTE by staff group, region, or organisation) but are not used for AP identification. These include ‘Staff Group and NHSE region’ (FTE and headcount by main staff group at region level), ‘Staff Group and Organisation’ (FTE and headcount by main staff group at organisation level), and ‘staff excluding medical’ (FTE by main staff group, care setting, level, and Agenda for Change band at organisation level). These files use the main staff group classification, not the NWD classification, and cannot identify Advanced Practitioners.

### Study population and Advanced Practitioner identification

The study population comprises all HCHS staff recorded in NHS Workforce Statistics with an AP-specific NWD Job Role code. Two distinct classification systems appear in the NHS Workforce Statistics and must not be conflated. The main staff group classification, used in the standard CSV files, groups staff by professional registration and NHS Occupation Code (for example, ‘Nurses & health visitors’, ‘Scientific, therapeutic & technical staff’). The NWD Staff Group and Job Role classification, used in Core 7 and Core 8, groups staff by a complementary system that includes specific Job Role codes such as ‘Nurse - Advanced Practitioner’ and ‘Paramedic Advanced Practitioner’. Only the NWD classification permits direct identification of Advanced Practitioners. Internal validation of the AP identification strategy was conducted by cross-referencing NWD Job Role codes against the NWD Staff Groups in which they are recorded, confirming that each AP code appeared only within the expected Staff Group (RECORD item 6.2).

The validation steps described above are internal to the published dataset. External validation, for instance comparing the published AP FTE figures against trust-level ESR extracts, organisational AP registers maintained by workforce leads, or direct confirmation from employing organisations, was considered but is not feasible within the constraints of this study design. Core 7 and Core 8 are pre-aggregated national publications; the underlying individual-level ESR records from which they are derived are not publicly available and would require a formal data access agreement with NHS England. The published data, therefore, cannot be linked to individual staff records, and no mechanism exists within this analytical framework to verify, at the record level, whether a given entry coded as an Advanced Practitioner meets the criteria set out in the Multi-Professional Framework. The internal cross-referencing of NWD Job Role codes against NWD Staff Group membership, together with the Core 7 to Core 8 cross-validation at the April 2026 time point, represents the maximum validation achievable with published aggregate data. The residual coding accuracy risk is addressed transparently in the Prospective limitations section below.

The primary analysis dataset will include qualified Advanced Practitioners only. Trainee Advanced Practitioners will be excluded from the primary analysis and will be presented separately as a clearly labelled supplementary sensitivity analysis. This separation is justified on both conceptual and methodological grounds. Qualified APs represent the workforce currently deployed at advanced practice level. Trainee APs are working towards but have not yet achieved the level of practice described in the MPF [2]. Combining the two groups would conflate current workforce capacity with future workforce supply. The trainee codes were also introduced from NWD v3.3 (February 2022) onwards and are therefore available for a shorter portion of the study period, which would complicate longitudinal trend interpretation.

The unit of analysis differs between the two analytical components, in accordance with STROSA-2 item 11. For the longitudinal analysis (Core 7), the unit is the published national-level FTE value at each combination of NWD Staff Group, NWD Job Role, Area of Work, and time point; trend tests operate on aggregated national time series with n = 13 time points per series. For the cross-sectional geographic analysis (Core 8), the primary unit is the NHS organisation (Trust or core organisation reporting AP FTE at April 2026); organisation-level FTE is then aggregated to NHS England Region (n = 7) and ICS (n = 42) for inequality and distributional analyses. Based on the structure of NHS England’s published organisational reporting, approximately 200 to 230 organisations are expected to contribute to the geographic analysis.

The full set of qualified AP Job Role codes used for programmatic filtering, confirmed by inspection of the February 2026 Core 7 and Core 8 data files during feasibility assessment, is presented in Table 2; the code list will be reconfirmed against the April 2026 release, including any NWD classification changes, prior to analysis. Four Trainee Advanced Practitioner codes were introduced at NWD v3.3, one within each of the NWD Staff Groups that carry qualified AP codes: Nursing and Midwifery Registered, Allied Health Professions, Additional Professional Scientific and Technical (APST), and Healthcare Scientists. These form the basis of the supplementary trainee analysis.

**Table 2.**
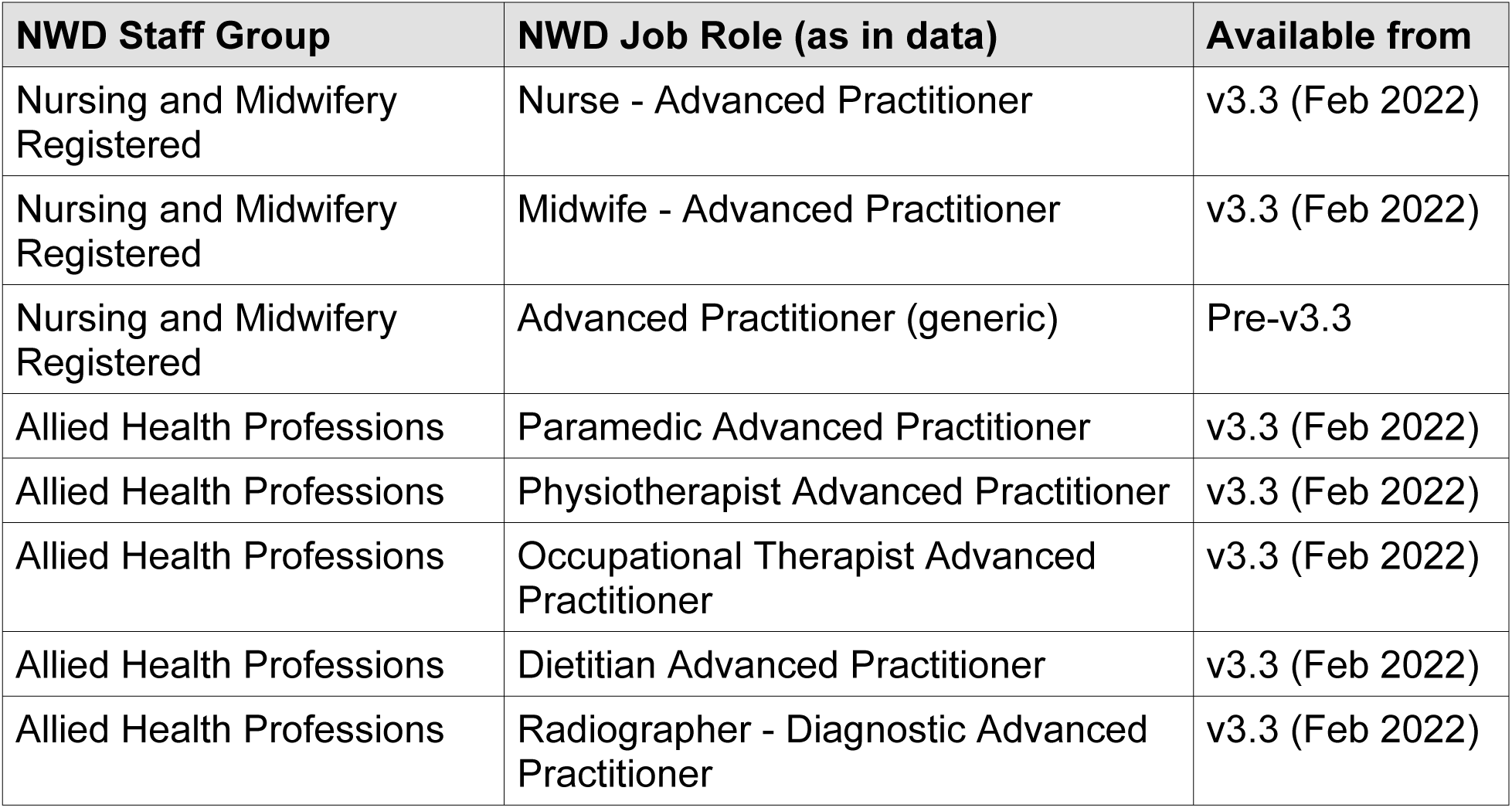
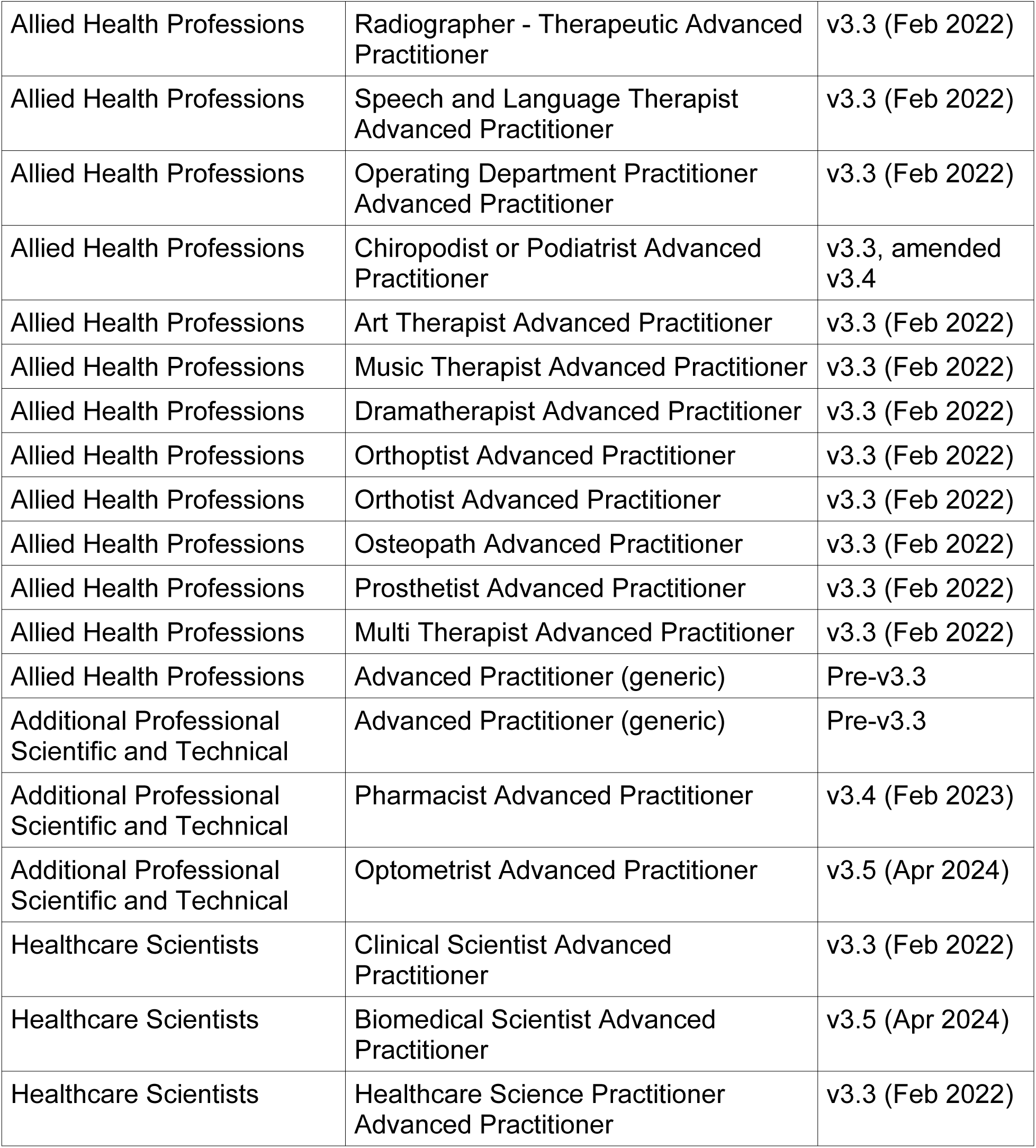
Qualified Advanced Practitioner NWD Job Role codes used for AP identification.

At the February 2026 feasibility inspection, 516 FTE remained coded under generic ‘Advanced Practitioner’ codes rather than the profession-specific codes introduced from v3.3 onwards (441 FTE in APST, 63 FTE in Nursing and Midwifery Registered, 11 FTE in Allied Health Professions). This indicates that not all trusts have migrated staff onto the newer codes. These generic codes are included in all analyses; they represent qualified Advanced Practitioners whose professional background cannot be determined from the published data. The persistence of generic coding is tracked as a sensitivity analysis.

### Study period

The longitudinal analysis uses all available time points in Core 7 from September 2014 to April 2026. September 2014 was selected as the start date because it is the first time point at which AP Job Role codes carry meaningful FTE data across multiple staff groups: 119 AP rows appeared in September 2014, compared with only 11 in September 2013. The series comprises annual September snapshots (2014 to 2025) plus the April 2026 endpoint, yielding 13 time points. While the September snapshots provide annual consistency, the April 2026 endpoint introduces a minor seasonal inconsistency that is noted as a limitation but is expected to have minimal effect on workforce composition data.

The study period spans three distinct phases in the development of the AP workforce and its measurement. The first phase (pre-MPF, September 2014 to September 2016) covers the period during which generic AP Job Role codes existed within the NWD, but no national AP framework had been published. The second phase (post-MPF, pre-classification expansion, September 2017 to September 2021) covers the period during which the MPF [1] was in effect, but the NWD continued to use only generic AP codes without profession-specific disaggregation. The third phase (post-NWD v3.3, September 2022 to April 2026) covers the period during which profession-specific AP Job Role codes were progressively introduced across NWD versions 3.3 to 3.6. These phases are used as descriptive context and annotated on trend figures to allow visual assessment of whether workforce trajectories change in relation to policy and classification milestones. They are not treated as formal analytical strata.

The geographic distribution analysis uses the April 2026 Core 8 file as a single cross-sectional snapshot. This is the only available time point at which organisation-level data with the NWD Job Role classification is published. Geographic inequality can therefore be described at one point in time but trends in geographic distribution cannot be assessed.

The progressive expansion of AP-specific Job Role codes across successive NWD versions affects the interpretation of longitudinal trends because new codes appearing in later versions April produce apparent workforce growth that partly reflects improved classification rather than genuine workforce expansion. Table 3 summarises the NWD classification version timeline and the AP-relevant changes at each version.

**Table 3.** NWD classification version timeline and AP-relevant changes.

| NWD version | Effective date | AP-relevant changes |
| --- | --- | --- |
| Pre-v3.3 | Pre-Feb 2022 | Generic AP codes in Nursing and Midwifery Registered, Allied Health Professions, and APST. No profession-specific disaggregation. |
| v3.3 | February 2022 | Profession-specific AP codes introduced for nursing, midwifery, and 15 allied health professions. Trainee AP codes introduced for all four Staff Groups. Clinical Scientist AP and Healthcare Science Practitioner AP introduced within Healthcare Scientists. |
| v3.4 | February 2023 | Pharmacist AP added within APST. Chiropodist or Podiatrist AP code amended. |
| v3.5 | April 2024 | Optometrist AP added within APST. Biomedical Scientist AP added within Healthcare Scientists. |
| v3.6 | February 2025 | No new AP codes. Enhanced Practice Nurse introduced within Nursing and Midwifery Registered (distinct from AP; excluded from this study). Area of Work renaming requiring harmonisation for longitudinal analysis. |

### Variables and classification systems

The primary outcome variable is full-time equivalent (FTE). In NHS Workforce Statistics, 1.0 FTE equates to 37.5 contracted hours per week. FTE is used in preference to headcount throughout because it provides an additive measure of workforce capacity that can be summed across organisations, staff groups, and areas of work without the double-counting risk inherent in headcount data (a single individual holding part-time positions in multiple organisations contributes to FTE in each but cannot be identified or deduplicated from published aggregate data). Cores 7 and 8 report FTE only; headcount is not available in these files.

All non-FTE variables function as stratifying or grouping variables rather than as predictors in a causal or predictive model. This terminology reflects the descriptive analytical intent of the study [26]. The stratifying variables are as follows. NWD Staff Group is a four-level categorical variable comprising Nursing and Midwifery Registered, Allied Health Professions, Additional Professional Scientific and Technical (APST), and Healthcare Scientists. NWD Job Role is nominal, with multiple AP-relevant codes (both generic and profession-specific) within each Staff Group, as detailed in Table 3. Area of Work is hierarchical, with primary, secondary, and tertiary levels defined by the NWD specification; it describes the clinical specialty or service area in which staff are deployed. NHS England Region is nominal with seven levels. ICS is nominal with 42 levels at 1 April 2026. Organisation is nominal, comprising all NHS Trusts and core organisations reporting AP FTE in Core 8. Time point takes 13 values (annual September 2014 to 2025 plus April 2026) and is treated as ordinal for trend tests. NWD version period is nominal with three levels (pre-MPF, post-MPF pre-classification expansion, post-NWD v3.3) and is used as figure annotation and descriptive contextualisation rather than as an analytical variable.

Derived summary measures computed from FTE include total qualified AP FTE (national and within strata), proportional distribution of AP FTE across strata with Wilson score 95% confidence intervals [33], compound annual growth rate (CAGR) as the headline endpoint-based growth metric, Mann-Kendall τ and Sen’s slope for trend characterisation, Kruskal-Wallis H with epsilon-squared effect size for cross-regional comparison, Gini coefficient for geographic inequality, and Dunn’s test statistics for pairwise regional comparisons. These are described in detail in the Statistical analysis plan (see below). No composite scales, indices, or derived constructs requiring item aggregation are used. No factor analysis is performed and no latent variables are estimated.

### Data preparation and quality assurance

The February 2026 Core 7 and Core 8 CSV files were downloaded from the NHS England NHS Workforce Statistics publication page on 1 April 2026 for feasibility inspection. The April 2026 release was selected as the analytical dataset because it is the most recent NHS Workforce Statistics publication available at the start of the analytical phase. Analysing the latest available release ensures that the longitudinal series and the cross-sectional geographic snapshot reflect the most current workforce position obtainable from the published data. The April 2026 Core 7 and Core 8 files will be downloaded from the same source following their scheduled release on 25 June 2026. Original files will be retained as an archival reference. Initial data cleaning and inspection will be performed in Microsoft 365 Excel for enterprise using Claude (Anthropic), an artificial intelligence (AI) assistant embedded within the Excel application, to support data preparation tasks. The Excel version to be used is 2605 (Build 20026.20182); the Claude model will be Opus 4.8. Should newer versions of either tool become available during the analytical phase, the versions used will be updated accordingly and the exact versions reported in the findings manuscript. All AI-assisted cleaning operations will be verified and approved by the lead author before the cleaned data are exported for analysis, in accordance with PLOS ONE editorial policy on the use of AI in research. The verified, cleaned datasets will then be imported into RStudio for all subsequent analysis in R. The AI tool will function as an assistant to the research team, not as an independent agent. All methodological decisions, including the choice of study design, the selection and justification of statistical tests, the specification of sensitivity analyses, the interpretation of preliminary data inspection findings, and the framing of the analytical approach within the descriptive epidemiology framework, were made by the authors. All statistical interpretations, inferential judgments, and analytical decisions reported in this protocol reflect the authors’ own reasoning.

Preliminary structural inspection of the February 2026 data files, conducted prior to protocol registration to verify data availability and inform the analytical design, confirmed the column structures described in the Data source and provenance section above. Date formatting differs between the two files: Core 7 uses YYYY-MM-DD format (for example, 2022-09-30) while Core 8 uses DD/MM/YYYY format (for example, 28/02/2026). Date formats will be standardised during data preparation.

Advanced Practitioners will be identified by extracting rows containing AP-specific Job Role codes from Core 7 and Core 8 using exact string matching on the NWD_JOB_ROLE variable. The complete list of qualified AP Job Role code strings, verified against the actual data values in the February 2026 files (Table 2), will be applied as an inclusion filter. A separate extraction using the four Trainee Advanced Practitioner Job Role codes (one per NWD Staff Group) will produce the trainee dataset for the supplementary analysis.

NHS Workforce Statistics apply small number suppression to protect individual confidentiality. Cells containing very small values are replaced with an asterisk in the published data. Inspection of the February 2026 Core 8 AP rows during feasibility assessment identified no suppressed values in the FTE column, and all FTE values were numeric. The April 2026 analytical files will be checked for suppression on release. Suppression handling will be reassessed if earlier time points in Core 7 contain suppressed cells. Suppressed values will be treated as missing data and reported transparently; they will not be imputed. Where suppressed values occur in cells contributing to derived totals, the derived total will be computed from the available non-suppressed values, and the number and location of suppressed cells will be documented.

Core 7 does not include calculated totals; these must be derived by summing the constituent FTE values. Where possible, derived national totals from Core 7 will be cross-validated against Core 8 by aggregating the April 2026 organisation-level data to national level and comparing the result. Any discrepancies will be documented and investigated.

NWD v3.6 (February 2025) introduced Area of Work renamings, including the renaming of Audiological Science to Audiology and Hearing Services with new adult and paediatric tertiary subdivisions, and the rehyphenation of several maxillofacial-related Area of Work values. These renamings require harmonisation during data preparation to ensure consistent longitudinal comparison across the study period. No statistical outliers will be removed. The right-skewed distribution of organisation-level FTE reflects the structural composition of the NHS provider sector, in which a small number of large acute trusts generate FTE values substantially higher than community, mental health, or specialist trusts. These are features of the workforce being characterised, not statistical artefacts.

### Statistical analysis plan

#### Framing and inferential justification

The NHS Workforce Statistics are a near-complete census of HCHS staff at each time point. The superpopulation framework provides the theoretical justification for

applying inferential statistics to census-type administrative data [34–38]. Under this framework, the observed workforce at each time point is treated as one realisation of an underlying stochastic data-generating process, driven by recruitment, retention, attrition, reclassification, and policy interventions, rather than as the exhaustive population of interest. This permits the use of confidence intervals and hypothesis tests, interpreted as reflecting the variability of the underlying workforce process rather than sampling error [39,40].

The appropriateness of inferential statistics on census data is contested. Gorard [41] argues that significance tests and confidence intervals are irrelevant when the full population of cases is used, since there is no sampling variation; Pollet [42] reaches a similar conclusion for macro-level population data. The superpopulation justification adopted here follows Berk, Western and Weiss [36] and Alexander [37] in treating the observed data as one realisation of a broader process, but acknowledges that this framing depends on the substantive plausibility of the data-generating mechanism described above. Consistent with this contested status, the study prioritises effect sizes and confidence intervals as the primary basis for substantive interpretation and de-emphasises p-values, which are reported for transparency but are not the basis for inferential claims about the AP workforce.

#### Descriptive statistics

Descriptive statistics form the core of the analysis. For each time point in the Core 7 series, the following will be reported: total AP FTE at national level; AP FTE by NWD Staff Group; AP FTE by Area of Work at primary, secondary, and tertiary levels of the NWD hierarchy; and proportional distribution across staff groups and areas of work. Where appropriate, these will be expressed as both absolute FTE values and as proportions of the relevant workforce denominator. Wilson score confidence intervals [33] will be used for all proportions. The Wilson score interval was selected in preference to the Wald (normal approximation) interval because it provides more accurate coverage, particularly for proportions near zero or one and for small denominators [43]. This is relevant because some AP staff group and Area of Work proportions April be very small, conditions under which the Wald interval produces intervals that extend below zero or above one and undercover the true proportion.

#### Longitudinal trend analysis

Change across the 13 time points will be described using absolute change (FTE difference), percentage change, and compound annual growth rate (CAGR) where the trend is approximately monotonic. CAGR is retained as the accessible headline growth metric for cross-report comparability with OECD [27], WHO [13], and Health Foundation workforce publications, but should be interpreted alongside the trend test results rather than in isolation, since CAGR uses only the endpoints and cannot detect change-points within the series. Trends will be presented graphically for key variables. The three-phase structure of the study period will be annotated on trend figures to allow visual assessment of whether workforce trajectories change in relation to these policy and classification milestones.

The monotonic trend in AP FTE will be formally tested using the Mann-Kendall trend test [44,45], a nonparametric test for monotonic trend that does not assume normality, linearity, or constant variance. Mann-Kendall was selected in preference to parametric alternatives (such as linear regression on time) because it makes no distributional assumptions, is valid for time series as short as n = 4, and tests directly for monotonic trend rather than imposing a linear functional form [46]. Where a significant monotonic trend is detected at α = 0.05, Sen’s slope estimator [47] will provide a robust estimate of the median annual rate of change in FTE. Sen’s slope was selected over the ordinary least squares regression slope because it is resistant to outliers and anomalous endpoint values [46], a property that matters here because the April 2026 endpoint is seasonally inconsistent with the annual September snapshots that comprise the remainder of the series. The Mann-Kendall test and Sen’s slope will be applied to the overall AP FTE series and to each NWD Staff Group individually. All tests are two-tailed; the descriptive framing does not pre-specify the direction of effects. No formal time series modelling (such as Autoregressive Integrated Moving Average (ARIMA)) will be applied, as 13 time points provide insufficient degrees of freedom for complex autoregressive approaches and the descriptive purpose of the study does not require causal modelling.

#### Cross-sectional geographic analysis

Geographic distribution of the AP workforce will be examined at NHS England region and ICS level using the April 2026 Core 8 data. Organisation-level FTE will be aggregated to region and ICS level using the NHSE_REGION_CODE and ICS_CODE identifiers embedded in the Core 8 file.

The Kruskal-Wallis test [48] will assess whether AP FTE distribution varies across the seven NHS England regions. Kruskal-Wallis is appropriate here because organisation-level FTE distributions are markedly right-skewed and heteroscedastic, conditions under which parametric Analysis of Variance (ANOVA) inflates Type I error. The test is interpreted as a test of stochastic dominance, that is, whether one region’s FTE distribution tends to produce systematically higher or lower values than another’s, rather than as a test of medians, consistent with the methodological guidance of Hart [49] and Divine et al. [50]. An effect size will be reported using epsilon-squared (ε² = H/((n²−1)/(n+1))), which ranges from 0 to 1. Epsilon-squared was selected as the effect size measure for the Kruskal-Wallis test because it is the non-parametric analogue of eta-squared and is directly interpretable as the proportion of variance in ranks accounted for by group membership [51]. Bias-corrected and accelerated (BCa) bootstrap 95% confidence intervals (2,000 resamples) will be reported for epsilon-squared. The BCa bootstrap was selected over percentile or standard bootstrap methods because it corrects for both bias and skewness in the bootstrap distribution [52], properties that are relevant where the sampling distribution of the effect size April be asymmetric, as is expected with right-skewed organisation-level FTE data. This addresses the concern that, with large n, even trivially small distributional differences can yield statistically significant Kruskal-Wallis results; the effect size quantifies the practical magnitude of regional variation. Where the Kruskal-Wallis omnibus is significant, Dunn’s test [53] will be applied as a post-hoc pairwise comparison across all 21 region pairs, re-using the global ranking from the omnibus test. Dunn’s test is methodologically preferable to pairwise Wilcoxon tests that re-rank within each pair [54].

The Gini coefficient [55] will summarise the degree of inequality in AP distribution across organisations and regions. Gini is the dominant inequality measure in the global health workforce equity literature [56,57] and its scale invariance, population-size independence, and satisfaction of the Pigou-Dalton transfer principle make it well suited to workforce distribution analysis. Bootstrap 95% confidence intervals (bias-corrected and accelerated, 2,000 resamples) will be calculated for all Gini estimates, following the recommendation of Dixon et al. [58] and Mills and Zandvakili [59]. The BCa method is preferred because the sampling distribution of the Gini coefficient is typically right-skewed, particularly at small sample sizes, and BCa corrects for both bias and skewness in the bootstrap distribution [52]. The small-sample correction factor n/(n−1) described by Deltas [60] will be applied when computing the Gini at region level (k = 7). A Lorenz curve will accompany every reported Gini to show where in the distribution the inequality is concentrated.

#### Multiple testing correction

The Benjamini-Hochberg procedure [61] will be applied to the 21 Dunn’s post-hoc pairwise comparisons and to any additional subgroup analyses. The Benjamini-Hochberg procedure controls the false discovery rate, the expected proportion of false positives among rejected hypotheses, which is more appropriate than family-wise error rate control (Bonferroni) for the descriptive screening aim of identifying which regions or subgroups warrant further investigation [62]. No global family-wise correction is applied across the full set of pre-specified tests, because the tests address distinct research questions on distinct data structures (longitudinal national time series versus cross-sectional organisation-level distributions) rather than constituting a single inferential family. This decision is consistent with the guidance of Rubin [63] against blanket multiplicity correction across substantively distinct hypotheses. Both unadjusted p-values and Benjamini-Hochberg-adjusted q-values will be reported to permit readers to apply alternative correction criteria.

#### Tests not included

Chi-square tests were excluded because the primary outcome (FTE) is continuous; binning continuous FTE into categories for chi-square analysis would involve an arbitrary and information-losing transformation not justified by the research questions. Linear regression and multiple linear regression were excluded because the study has a descriptive aim with no pre-specified causal estimand or theoretically motivated predictors [26], the data are a census rather than a sample, and the 13 aggregate time points provide insufficient degrees of freedom for stable multivariable estimation. This descriptive approach is consistent with the conventions of major workforce reporting bodies including the OECD [27], WHO [13], the Health Foundation [28], and the Nuffield Trust [29,30], and with the descriptive epidemiology framework of Lesko, Fox and Edwards [25] and Fox et al. [64].

#### Inference criteria

Inferences will be drawn from the joint pattern of estimated magnitudes, confidence intervals, effect sizes, and (where applicable) p-values, with substantively meaningful magnitudes prioritised over statistical significance alone. The significance threshold is α = 0.05 where p-values are reported, two-tailed throughout. All point estimates of proportions, growth rates, slopes, effect sizes, and inequality measures are reported with 95% confidence intervals: Wilson score for proportions, BCa bootstrap (2,000 resamples) for epsilon-squared and Gini, and standard confidence intervals for Sen’s slope. Where p-values and effect sizes diverge (for example, a Kruskal-Wallis test significant at p < 0.001 with epsilon-squared < 0.01), the effect size and confidence interval will be the primary basis for substantive interpretation, and the p-value will be reported but explicitly de-emphasised.

### Sensitivity analyses

Six pre-specified sensitivity analyses will be conducted to test the robustness of the primary findings to key methodological decisions and data quality constraints.

The primary analysis will be repeated with Trainee Advanced Practitioners included alongside qualified APs. At the February 2026 feasibility inspection, trainees accounted for 2,023 FTE nationally. The trainee population will also be described separately for workforce pipeline analysis (research question 5). A ‘generic codes only’ sensitivity analysis will recompute the national AP FTE trend using only the pre-v3.3 generic AP Job Role codes across the full study period, providing a like-for-like trend uncontaminated by classification expansion. A separate ‘constant code set’ analysis will compute the trend from September 2022 (NWD v3.3) onwards using only those AP codes present from v3.3, testing whether apparent growth within the post-classification-expansion period is driven by genuine workforce expansion or by the progressive introduction of new codes in v3.4 and v3.5.

The proportion of AP FTE coded under generic ‘Advanced Practitioner’ codes rather than profession-specific codes will be tracked across post-v3.3 time points. A declining proportion indicates progressive migration to the newer codes; a stable proportion suggests persistent coding inconsistency across trusts. The Gini coefficient will be reported in two forms: unweighted (each organisation treated as a unit) and weighted by total HCHS FTE at organisation level, to characterise whether observed geographic inequality reflects the distribution of AP roles per se or the distribution of organisations of varying size. The modified Mann-Kendall test with variance correction for serial dependence [65] will be reported alongside the standard test for every series in which a significant trend is detected, and the Kruskal-Wallis and Gini analyses will be repeated within stratified subsets (for example, acute trusts only; community and mental health trusts only) to characterise whether regional variation persists after accounting for organisational type.

The non-parametric and rank-based methods selected for this study were chosen specifically because they do not assume normality, homoscedasticity, or large-sample asymptotic behaviour, and they tolerate the right-skewed and heteroscedastic distributions characteristic of organisation-level NHS FTE data. The principal residual assumption is that of independent observations within each test, which is plausible at organisation level (each Trust is an administratively distinct reporting unit) but is necessarily relaxed for the longitudinal national time series, where autocorrelation between adjacent annual snapshots is expected. The Mann-Kendall test is robust to serial dependence at the magnitudes typically observed in annual workforce series; the modified Mann-Kendall sensitivity analysis addresses this directly. Bootstrap procedures will use 2,000 BCa resamples. If any bootstrap fails to converge, the failure will be reported transparently, an alternative confidence interval method (percentile bootstrap or analytic confidence interval) will be substituted, and the substitution will be documented.

### Software and reproducibility

Initial data cleaning and inspection will be performed in Microsoft Excel with Claude (Anthropic) as an AI-assisted data preparation tool, with all operations verified by the lead author. All subsequent data preparation and statistical analysis will be conducted in R (version 4.6.1, ‘Happy Hop’) [66] using RStudio (version 2026.05.1) as the integrated development environment. The tidyverse collection of packages will be used for data manipulation and visualisation, including dplyr for data wrangling, ggplot2 for figures, and readr for file import. The DescTools package will provide Wilson score confidence intervals and the Gini coefficient with small-sample correction. The stats package (included in base R) provides the Kruskal-Wallis test via kruskal.test and the Benjamini-Hochberg correction via p.adjust. The FSA package will provide Dunn’s post-hoc test via dunnTest. The rstatix package will provide the Kruskal-Wallis effect size (epsilon-squared) via kruskal_effsize. The Kendall package will provide the Mann-Kendall trend test and Sen’s slope estimator. The boot package (included in base R) will be used for bootstrap confidence intervals for the Gini coefficient. The ineq package will be used for Lorenz curve construction.

All analysis code will be made available as a supplementary file on publication. The underlying NHS Workforce Statistics data are publicly available from the NHS England website.

### Ethical considerations

This study uses publicly available, aggregate-level administrative data published under the Open Government Licence v3.0. No individual-level data are accessed.

The study has been registered with Northumbria University (ethics ID: 2025-10406-11580). Ethical approval via NHS Research and Ethics Committee for data access is not required for secondary analysis of published aggregate data, consistent with Health Research Authority guidance.

## Discussion

### Prospective strengths

The MAPPED study will provide the first national, longitudinal quantification of the Advanced Practitioner workforce in NHS Hospital and Community Health Services in England derived from routinely collected administrative data. No published analysis has previously used the NHS Workforce Statistics to characterise this workforce across professional groups, clinical specialties, and geographic areas over time. The study addresses a gap that has persisted despite the policy prominence of the AP role and the availability of publicly accessible workforce data containing the relevant classification codes.

A principal advantage of the secondary data approach is coverage. The NHS Workforce Statistics are a near-complete census of HCHS staff at each publication time point, derived from validated Electronic Staff Record extracts covering all NHS Trusts and core organisations in England. No primary data collection method, whether survey, audit, or registration-based, could achieve equivalent national coverage at comparable cost. Survey-based approaches to AP workforce enumeration, while valuable for capturing scope of practice and qualitative workforce characteristics, are subject to response bias and cannot produce the population-level denominators required for workforce planning. The Fothergill et al. [5] national survey of over 4,000 AP staff, for instance, provided important evidence on role variation and governance but was not designed as a geographic enumeration and did not report region-level workforce counts. The Lawler et al. [6] mixed-methods evaluation included respondents from different English regions but was similarly not structured for national quantification. The MAPPED study uses a reproducible data source that is publicly available under the Open Government Licence v3.0, enabling independent verification and replication.

Few countries have managed to count their advanced practice workforce reliably from routine data. A global survey of 325 respondents across 26 countries reported that understanding the number, distribution, and types of advanced practice providers is “extremely complicated” because of categorisation issues specific to nursing and advanced practice [22]. A six-country analysis of nurse practitioner workforce size excluded UK nations because no suitable national registry or data source was identified [18]. In Australia, position titles alone were insufficient for AP classification, requiring the development of a bespoke role-delineation tool applied to a survey of 5,662 respondents [23]. In the United States, a validation study of the National Provider Identifier file found that it undercounted advanced practice midwives because multiple taxonomy categories existed for the same role [24]. The MAPPED study demonstrates that routinely collected administrative data, despite known coding limitations, can be used for national AP workforce enumeration where the classification system contains role-specific codes. This finding has potential relevance for other countries developing or expanding their administrative workforce data systems.

The longitudinal depth of the Core 7 time series (13 time points spanning September 2014 to April 2026) permits characterisation of AP workforce growth across more than a decade, encompassing both the period before and after the introduction of profession-specific NWD Job Role codes. This temporal range is substantially longer than existing survey-based snapshots, which typically capture a single time point or a limited window. Regional workforce reports from the East of England and the North West have documented AP growth within their geographies, but no published analysis has examined the national longitudinal trajectory. The three-phase periodisation enables visual assessment of whether growth trajectories change in relation to policy milestones (the 2017 MPF) and classification changes (the NWD v3.3 expansion from February 2022), though the study does not claim causal relationships between these events and workforce trends.

The analytical framework is designed to be transparent about its assumptions and limitations. The pre-specified analytical plan, registered on the Open Science Framework (OSF) prior to formal analysis, distinguishes clearly between pre-specified descriptive analyses and exploratory analyses. The superpopulation justification for applying inferential statistics to census data is explicitly stated and its contested status acknowledged, with effect sizes and confidence intervals prioritised over p-values. The six pre-specified sensitivity analyses directly address the principal threats to interpretive validity: classification expansion effects, generic code persistence, serial dependence in the trend series, organisational mix effects on regional comparisons, and the influence of Trainee AP inclusion. The study design also responds to an identified weakness in the published PLOS ONE secondary data analysis literature: a review of comparator protocol papers found inconsistent reporting guideline compliance, and the RECORD/STROSA approach adopted here positions this study as methodologically more rigorous in its reporting than much of the existing secondary analysis literature. The analysis code will be deposited as supplementary material alongside the findings paper for transparency and to enable further longitudinal study as further NWD becomes available.

### Prospective limitations

Five limitations of this study should be considered prospectively, each arising from the nature of the data source rather than the analytical design.

The accuracy of NWD Job Role classification depends on coding practice at trust level. The Centre for Advancing Practice ESR coding guidance [4] acknowledges that coding accuracy varies across organisations. Staff working at advanced practice level April not have the AP Job Role code recorded in ESR, particularly where trusts have not updated local coding following the introduction of profession-specific codes. Conversely, staff with ‘advanced’ in their job title who do not meet the MPF criteria April have the code applied. A secondary analysis of 17,960 specialist nursing posts identified 595 distinct job titles, with clustering driven primarily by pay band rather than education or competency [67], which illustrates the scale of title variation that ESR coding must navigate. The published data represent the workforce as coded, not necessarily as deployed. This is a structural constraint of secondary analysis of administrative data; it cannot be resolved without access to the underlying individual-level ESR records, which are not publicly available. External validation against trust-level data sources would strengthen confidence in the published figures but was not feasible here.

Organisational AP registers, where they exist, are maintained locally and there is no national repository or standardised format that would permit systematic comparison. Workforce leads in individual trusts could in principle confirm whether the AP-coded FTE reported in their ESR submission reflects actual AP deployment, but this would require a separate primary data collection exercise across the approximately 200 to 230 contributing organisations, which falls outside the scope of a secondary analysis of published statistics. Future research with access to linked, individual-level ESR data, or with targeted primary validation in a purposive sample of trusts, could quantify the gap between published AP FTE and the workforce as actually deployed. Until such validation is available, the figures reported in this study should be interpreted as the AP workforce as coded in ESR, which April undercount practitioners in organisations with incomplete coding and overcount in organisations that apply the AP Job Role code to staff who do not meet the MPF criteria.

Generic ‘Advanced Practitioner’ codes persist alongside the profession-specific codes (516 FTE at the February 2026 feasibility inspection), which means that the professional background of a proportion of the AP workforce cannot be determined from the published data. The majority of this generic coding is concentrated in the Additional Professional Scientific and Technical staff group (441 FTE), which April reflect slower uptake of the Pharmacist Advanced Practitioner code introduced at v3.4 or the presence of professions for which no specific code yet exists. Regional evidence confirms that coding adoption is incomplete: in the East of England, only 56% of secondary care organisations had updated job roles to the profession-specific codes at the time of a regional workforce report [21]. The generic code persistence sensitivity analysis will characterise the extent of this issue but cannot eliminate it.

The longitudinal analysis uses September snapshots for 2014 to 2025 and a April endpoint for 2026. While seasonal effects on workforce composition data are expected to be minimal (ESR records contracted FTE, which is less susceptible to seasonal variation than activity-based measures), this inconsistency is noted. More substantially, the transition from generic to profession-specific codes from NWD v3.3 (February 2022) onwards creates a measurement discontinuity within the time series. Regional reports have acknowledged that recorded AP growth April partly reflect improvements in ESR coding rather than genuine workforce expansion. The two classification version sensitivity analyses (‘generic codes only’ and ‘constant code set’) are designed to address this, but they cannot fully separate real growth from classification effects where both occurred simultaneously. This challenge is not unique to the MAPPED study; it is the central methodological problem facing any longitudinal analysis of a workforce category whose measurement system changed during the study period.

The geographic analysis is limited to a single cross-sectional snapshot (April 2026) because historical Core 8 files with the NWD classification are not published by NHS England. Geographic inequality in AP deployment can therefore be described at one point in time but trends in geographic distribution cannot be assessed. Whether the regional variation observed at April 2026 is widening, narrowing, or stable over time remains unknown. International evidence suggests that this question matters: in rural areas of the United States, nurse practitioner supply rose while physician supply fell between 2010 and 2016 [16], and a retrospective study of 1,255 Primary Care Networks in England found that new clinical roles introduced through the Additional Roles Reimbursement Scheme did not substantially change longstanding inequalities in workforce distribution [68]. Whether AP deployment in England follows a similar pattern of geographic persistence is an important question for future research.

FTE data do not capture the number of individual practitioners. A single person holding multiple part-time AP positions across organisations would contribute to FTE in each organisation but could not be identified or deduplicated in the published aggregate data. FTE is therefore a measure of deployed workforce capacity rather than a headcount of individual Advanced Practitioners. For workforce planning purposes, FTE is arguably the more relevant metric, since it directly reflects service capacity; but readers should be aware that the number of individuals working in AP roles April differ from the FTE figures reported.

### Implications for future research

The MAPPED study addresses one specific gap, the absence of national, longitudinal, routinely-data-derived AP workforce intelligence, but it sits within a broader set of unmet research needs that the findings paper cannot resolve alone.

The most immediate need is for consistent national operational definitions of AP roles that can be applied reliably in ESR coding. Despite the 2025 framework update, variation in how ‘advanced practitioner’ is understood and applied persists across organisations [5,7,9]. International experience suggests that alignment between regulatory bodies, employers, and data systems is a prerequisite for reliable enumeration [22]. Improving ESR coding accuracy is not merely a data quality exercise; it is a workforce planning prerequisite.

The geographic analysis in MAPPED, limited as it is to a single snapshot, highlights the need for routine publication of AP FTE at ICS and regional level, stratified by staff group and clinical area. The infrastructure for this exists within the monthly HCHS workforce statistics; what is needed is disaggregation by AP-specific Job Role codes. Moving beyond raw FTE to need-adjusted indicators (for example, AP FTE per weighted population) would enable more meaningful comparison across geographies and support targeted investment in underserved areas. International examples of supply-to-population ratios in rural workforce planning provide methodological precedents [16,69,70].

The trainee AP workforce, described in MAPPED as a supplementary analysis, warrants sustained longitudinal monitoring. Trainee starts are a necessary but insufficient indicator of future supply. Without progression and retention data linking trainee AP records to subsequent qualified AP coding in ESR, it is not possible to calibrate education commissioning against actual workforce output. A qualitative evidence synthesis of trainee AP role transition found that where orientation, mentorship, clinical supervision, and Master’s-level education were neglected, there was often failure to reach expected goals or resignation from the role [71]. The NHS Long Term Workforce Plan projects that over 6,300 clinicians will start advanced practice pathways annually by 2031/32 [3]; whether this training investment translates into qualified AP capacity in the specialties and geographies where it is most needed is a question that current data systems cannot yet answer.

Finally, the difficulties of international AP workforce comparison documented in the literature [18,19,22] point to a need for internationally comparable AP workforce indicators. The five-year review of the WHO Global Strategy found that most multilateral and bilateral agencies providing health workforce assistance did not identify human resources for health as a specific strategic focus, and few had published dedicated workforce investment policies [14]. England’s experience with multi-professional AP coding and national framework development could inform international efforts to improve comparability, while learning from countries with more established registry-based enumeration. A minimum common dataset for AP workforce reporting, respecting national differences in regulation and role design, would address one dimension of this gap and support the kind of cross-country learning that workforce planners in all participating countries would benefit from.

### Timeline and status

This paper presents the study protocol; no analysis of the planned dataset has been undertaken and no study results have been generated. As a secondary analysis of published aggregate data, the study involves no participant recruitment. A limited structural inspection of the February 2026 NHS Workforce Statistics release was conducted in April 2026 to confirm the data structure, variable availability, and feasibility, and to inform the analytical design; this inspection is disclosed under the prior knowledge statement in the OSF registration and did not constitute the study’s data collection. The analytical dataset is the April 2026 NHS Workforce Statistics release (Core 7 and Core 8), which NHS England is scheduled to publish on 26 June 2026 and which had not been released at the time of submission of this protocol. Data collection will be completed following that publication, when the April 2026 Core 7 and Core 8 files are downloaded. Data preparation and statistical analysis will be undertaken between 1 July and 31 August 2026, with results anticipated in early September 2026. The study protocol was registered as a Secondary Data Preregistration on the OSF on 18 April 2026 (doi: 10.17605/OSF.IO/DWSF3), prior to data collection and analysis; an update documenting the revised analytical endpoint and timeline was posted on 25 June 2026. The findings paper, reporting the results of the analyses described in this protocol, will be submitted for publication as a separate manuscript.

### Dissemination

The primary output of this study will be a findings paper reporting the results of the analyses described in this protocol, to be submitted for peer-reviewed publication. The underlying NHS Workforce Statistics data are publicly available from the NHS England website under the Open Government Licence v3.0. All analysis code will be deposited as a supplementary file alongside the findings paper, enabling independent verification and replication. The study protocol has been registered on the OSF. Findings will be presented at conferences and shared with NHS England’s Centre for Advancing Practice and relevant professional bodies to inform workforce planning and ESR coding policy.

### Supporting information

The following supporting information files will accompany this manuscript.

- **S1 Checklist. RECORD reporting checklist.**
- **S2 Table. STROSA supplementary items mapping.**

## Data Availability

No datasets were generated or analysed during the current study. All relevant data from this study will be made available upon study completion.

## Acknowledgments

The authors acknowledge NHS England (formerly NHS Digital) for the production and publication of the NHS Workforce Statistics used in this study.

## Author Contributions

**Conceptualisation:** Sadie Diamond-Fox, Barry Hill

**Data curation:** Sadie Diamond-Fox

**Formal analysis:** Sadie Diamond-Fox, Barry Hill

**Funding acquisition:** Not applicable

**Methodology:** Sadie Diamond-Fox, Barry Hill

**Project administration:** Sadie Diamond-Fox

**Resources:** Sadie Diamond-Fox

**Writing – original draft:** Sadie Diamond-Fox

**Writing – review & editing:** Sadie Diamond-Fox, Barry Hill

## Notes

### Competing Interest Statement

The authors have declared no competing interest.

### Funding Statement

The author(s) received no specific funding for this work.

### Author Declarations

The study has been registered with Northumbria University (ethics registration 2025-10406-11580) as it uses only publicly available, aggregate administrative data, formal ethical approval for data access is not required.

## References

1. Health Education England. Multi-professional framework for advanced clinical practice in England. London: HEE; 2017. Available from: https://www.hee.nhs.uk/sites/default/files/documents/multi-professionalframeworkforadvancedclinicalpracticeinengland.pdf

2. NHS England. Multi-professional framework for advanced practice in England. 2nd ed. London: NHS England; 2025. Available from: https://advanced-practice.hee.nhs.uk/multi-professional-framework-for-advanced-practice/

3. NHS England. NHS Long Term Workforce Plan. London: NHS England; 2023.

4. NHS England Centre for Advancing Practice. A guide to ESR coding for advanced practitioner roles, version 3. London: NHS England; 2025. Available from: https://advanced-practice.hee.nhs.uk/resources/a-guide-to-esr-coding-for-advanced-practitioner-roles/

5. Fothergill LJ, Al-Oraibi A, Houdmont J, Conway J, Evans C. Nationwide evaluation of the advanced clinical practitioner role in England: a cross-sectional survey. BMJ Open. 2022;12(1):e055475.

6. Lawler J, Maclaine K, Leary A. Workforce experience of the implementation of an advanced clinical practice framework in England: a mixed methods evaluation. Hum Resour Health. 2020;18(1):96.

7. Mann C, Timmons S, Evans C, Pearce R, Overton C, Hinsliff-Smith K, et al. Exploring the role of advanced clinical practitioners (ACPs) and their contribution to health services in England: a qualitative exploratory study. Nurse Educ Pract. 2023;67:103546. doi: 10.1016/j.nepr.2023.103546

8. Kuczawski M, Ablard S, Sampson F, Croft S, Sutton-Klein J, Mason S. Exploring advanced clinical practitioner perspectives on training, role identity and competence: a qualitative study. BMC Nurs. 2024;23(1):185. doi: 10.1186/s12912-024-01843-x

9. Timmons S, Mann C, Evans C, Pearce R, Overton C, Hinsliff-Smith K. The Advanced Clinical Practitioner (ACP) in UK healthcare: dichotomies in a new ‘multi-professional’ profession. SSM Qual Res Health. 2023;3:100211. doi: 10.1016/j.ssmqr.2022.100211

10. Evans C, Poku B, Pearce R, Eldridge J, Hendrick P, Knaggs R, et al. Characterising the outcomes, impacts and implementation challenges of advanced clinical practice roles in the UK: a scoping review. BMJ Open. 2021;11(8):e048171. doi: 10.1136/bmjopen-2020-048171

11. Buchan J, Campbell J. Challenges posed by the global crisis in the health workforce. BMJ. 2013;347:f6201.

12. Frenk J, Chen L, Bhutta ZA, Cohen J, Crisp N, Evans T, et al. Health professionals for a new century: transforming education to strengthen health systems in an interdependent world. Lancet. 2010;376(9756):1923–1958.

13. World Health Organization. Global strategy on human resources for health: workforce 2030. Geneva: WHO; 2016. Available from: https://www.who.int/publications/i/item/9789241511131

14. McIsaac M, Buchan J, Abu-Agla A, Kawar R, Campbell J. Global Strategy on Human Resources for Health: Workforce 2030, a five-year check-in. Hum Resour Health. 2024;22(1):68. doi: 10.1186/s12960-024-00940-x

15. Auerbach DI, Staiger DO, Buerhaus PI. Growing ranks of advanced practice clinicians: implications for the physician workforce. N Engl J Med. 2018;378(25):2358–2360.

16. Xue Y, Smith JA, Spetz J. Primary care nurse practitioners and physicians in low-income and rural areas, 2010-2016. JAMA. 2019;321(1):102–105.

17. Patel SY, Auerbach D, Huskamp HA, Frakt A, Neprash H, Barnett ML, et al. Provision of evaluation and management visits by nurse practitioners and physician assistants in the USA from 2013 to 2019: cross-sectional time series study. BMJ. 2023;382:e073933. doi: 10.1136/bmj-2022-073933

18. Maier CB, Barnes H, Aiken LH, Busse R. Descriptive, cross-country analysis of the nurse practitioner workforce in six countries: size, growth, physician substitution potential. BMJ Open. 2016;6:e011901.

19. Delamaire ML, Lafortune G. Nurses in advanced roles: a description and evaluation of experiences in 12 developed countries. OECD Health Working Papers No. 54. Paris: OECD Publishing; 2010. doi: 10.1787/5kmbrcfms5g7-en

20. NHS England. NHS Workforce Statistics methodology notes. London: NHS England; 2023. Available from: https://digital.nhs.uk/data-and-information/publications/statistical/nhs-workforce-statistics

21. NHS England East of England Faculty for Advancing Practice. Overview of the advanced practice workforce in the East of England 2023/24. London: NHS England; 2024. Available from: https://advanced-practice.hee.nhs.uk/wp-content/uploads/sites/28/2024/03/Overview-of-the-advanced-practice-workforce-in-the-East-of-England-.pdf

22. Wheeler KJ, Miller M, Pulcini J, Gray D, Ladd E, Rayens MK. Advanced practice nursing roles, regulation, education, and practice: a global study. Ann Glob Health. 2022;88(1):42. doi: 10.5334/aogh.3698

23. Gardner G, Duffield C, Doubrovsky A, Adams M. Identifying advanced practice: a national survey of a nursing workforce. Int J Nurs Stud. 2016;55:60–70.

24. Vanderlaan J, Jefferson K. Evaluation of a method to identify midwives in national provider identifier data. BMC Pregnancy Childbirth. 2023;23(1):809. doi: 10.1186/s12884-023-06122-2

25. Lesko CR, Fox MP, Edwards JK. A framework for descriptive epidemiology. Am J Epidemiol. 2022;191(12):2063–2070.

26. Hernán MA, Hsu J, Healy B. A second chance to get causal inference right: a classification of data science tasks. Chance. 2019;32(1):42–49.

27. Organisation for Economic Co-operation and Development. Health at a glance 2023: OECD indicators. Paris: OECD Publishing; 2023. Available from: https://www.oecd.org/en/publications/health-at-a-glance-2023_7a7afb35-en/full-report.html

28. Buchan J, Charlesworth A, Gershlick B, Seccombe I. A critical moment: NHS staffing trends, retention and attrition. London: The Health Foundation; 2019. Available from: https://www.health.org.uk/reports-and-analysis/reports/a-critical-moment

29. Julian S, Morris J, Scobie S. Independent prescribing in the UK: workforce ambitions and implementation challenges. London: Nuffield Trust; 2026. Available from: https://www.nuffieldtrust.org.uk/research/independent-prescribing-in-the-uk-workforce-ambitions-and-implementation-challenges

30. Palmer W, Crellin N, Lobont C. In the balance: lessons for changing the mix of professions in NHS services. London: Nuffield Trust; 2025. Available from: https://www.nuffieldtrust.org.uk/research/in-the-balance-lessons-for-changing-the-mix-of-professions-in-nhs-services

31. Benchimol EI, Smeeth L, Guttmann A, Harron K, Moher D, Petersen I, et al. The REporting of studies Conducted using Observational Routinely-collected health Data (RECORD) statement. PLoS Med. 2015;12(10):e1001885.

32. Swart E, Bitzer EM, Gothe H, Harling M, Hoffmann F, Horenkamp-Sonntag D, et al. A consensus German reporting standard for secondary data analyses, version 2 (STROSA-STandardisierte BerichtsROutine für SekundärdatenAnalysen). Gesundheitswesen. 2016;78(S 01):e145-e160. doi: 10.1055/s-0042-108647

33. Wilson EB. Probable inference, the law of succession, and statistical inference. J Am Stat Assoc. 1927;22(158):209–212.

34. Hartley HO, Sielken RL. A “super-population viewpoint” for finite population sampling. Biometrics. 1975;31(2):411–422.

35. Isaki CT, Fuller WA. Survey design under the regression superpopulation model. J Am Stat Assoc. 1982;77(377):89–96.

36. Berk RA, Western B, Weiss RE. Statistical inference for apparent populations. Sociol Methodol. 1995;25:421–458.

37. Alexander N. What’s more general than a whole population? Emerg Themes Epidemiol. 2015;12:11. doi: 10.1186/s12982-015-0029-4

38. Little RJ. To model or not to model? Competing modes of inference for finite population sampling. J Am Stat Assoc. 2004;99(466):546–556. doi: 10.1198/016214504000000467

39. Graubard BI, Korn EL. Inference for superpopulation parameters using sample surveys. Stat Sci. 2002;17(1):73–96.

40. Gibbs BG, Shafer K, Dufur MJ. Why infer? The use and misuse of population data in sport research. Int Rev Sociol Sport. 2015;50(1):115–121.

41. Gorard S. Do we really need confidence intervals in the new statistics? Int J Soc Res Methodol. 2019;22(3):281–291. doi: 10.1080/13645579.2018.1525064

42. Pollet TV. Much ado about p. What does a p-value mean when testing hypotheses with aggregated cross-cultural data in the field of evolution and human behavior? Front Psychol. 2013;4:734.

43. Newcombe RG. Two-sided confidence intervals for the single proportion: comparison of seven methods. Stat Med. 1998;17(8):857–872.

44. Mann HB. Nonparametric tests against trend. Econometrica. 1945;13(3):245–259.

45. Kendall MG. Rank correlation methods. 4th ed. London: Charles Griffin; 1975.

46. Helsel DR, Hirsch RM. Statistical methods in water resources. Techniques of Water-Resources Investigations, Book 4, Chapter A3. Reston (VA): US Geological Survey; 2002.

47. Sen PK. Estimates of the regression coefficient based on Kendall’s tau. J Am Stat Assoc. 1968;63(324):1379–1389.

48. Kruskal WH, Wallis WA. Use of ranks in one-criterion variance analysis. J Am Stat Assoc. 1952;47(260):583–621.

49. Hart A. Mann-Whitney test is not just a test of medians: differences in spread can be important. BMJ. 2001;323(7309):391–393.

50. Divine GW, Norton HJ, Barón AE, Juarez-Colunga E. The Wilcoxon-Mann-Whitney procedure fails as a test of medians. Am Stat. 2018;72(3):278–286.

51. Tomczak M, Tomczak E. The need to report effect size estimates revisited: an overview of some recommended measures of effect size. Trends Sport Sci. 2014;1(21):19–25.

52. Efron B. Better bootstrap confidence intervals. J Am Stat Assoc. 1987;82(397):171–185.

53. Dunn OJ. Multiple comparisons using rank sums. Technometrics. 1964;6(3):241–252.

54. Midway S, Robertson M, Flinn S, Kaller M. Comparing multiple comparisons: practical guidance for choosing the best multiple comparisons test. PeerJ. 2020;8:e10387. doi: 10.7717/peerj.10387

55. Gini C. Variabilità e mutabilità. Bologna: C. Cuppini; 1912.

56. Anand S. Measuring health workforce inequalities: methods and application to China and India. Human Resources for Health Observer No. 5. Geneva: WHO; 2010.

57. Brown MC. Using Gini-style indices to evaluate the spatial patterns of health practitioners: theoretical considerations and an application based on Alberta data. Soc Sci Med. 1994;38(9):1243–1256.

58. Dixon PM, Weiner J, Mitchell-Olds T, Woodley R. Bootstrapping the Gini coefficient of inequality. Ecology. 1987;68(5):1548–1551.

59. Mills JA, Zandvakili A. Statistical inference via bootstrapping for measures of inequality. J Appl Econom. 1997;12(2):133–150.

60. Deltas G. The small-sample bias of the Gini coefficient: results and implications for empirical research. Rev Econ Stat. 2003;85(1):226–234.

61. Benjamini Y, Hochberg Y. Controlling the false discovery rate: a practical and powerful approach to multiple testing. J R Stat Soc Series B Stat Methodol. 1995;57(1):289–300.

62. Glickman ME, Rao SR, Schultz MR. False discovery rate control is a recommended alternative to Bonferroni-type adjustments in health studies. J Clin Epidemiol. 2014;67(8):850–857.

63. Rubin M. When to adjust alpha during multiple testing: a consideration of disjunction, conjunction, and individual testing. Synthese. 2021;199:10969–11000.

64. Fox MP, Murray EJ, Lesko CR, Sealy-Jefferson S. On the need to revitalize descriptive epidemiology. Am J Epidemiol. 2022;191(7):1174–1179.

65. Hamed KH, Rao AR. A modified Mann-Kendall trend test for autocorrelated data. J Hydrol. 1998;204(1-4):182–196.

66. R Core Team. R: a language and environment for statistical computing. Vienna: R Foundation for Statistical Computing; 2026. Available from: https://www.R-project.org/

67. Leary A, Maclaine K, Trevatt P, Radford M, Punshon G. Variation in job titles within the nursing workforce. J Clin Nurs. 2017;26(23-24):4945–4950.

68. Hutchinson J, Lau YS, Sutton M, Checkland K. How new clinical roles in primary care impact on equitable distribution of workforce: a retrospective study. Br J Gen Pract. 2023;73(734):e659–e666. doi: 10.3399/BJGP.2023.0007

69. Hecht CJ 2nd, Burkhart RJ, McNassor R, Acuña AJ, Kamath AF. What is the geographic distribution and density of orthopaedic advanced practice professionals in rural counties? A large-database study. Clin Orthop Relat Res. 2023;481(10):1907–1916. doi: 10.1097/CORR.0000000000002649

70. Argus G, Gavaghan B, Spinks J. Health workforce planning methods in rural and remote primary care: a scoping review. Hum Resour Health. 2026;24(1):19. doi: 10.1186/s12960-026-01060-4

71. Moran GM, Nairn S. How does role transition affect the experience of trainee Advanced Clinical Practitioners: qualitative evidence synthesis. J Adv Nurs. 2018;74(2):251–262.

